# Endometriosis risk is associated with shorter anogenital distance by meta-analysis

**DOI:** 10.1101/2024.01.05.24300901

**Authors:** Bernard J. Crespi

## Abstract

**Background:** Anogenital distance is a well-validated marker of prenatal testosterone, with shorter distances indicating lower levels during early gestation. A suite of studies has linked anogenital distance with risk of endometriosis, but the findings are variable, leading to uncertainty in interpretation. The relationship of anogenital distance with endometriosis is especially important because lower testosterone has been associated with endometriosis in recent Mendelian Randomization studies, which implies causality in the association, with direct implications for future research and treatment.

**Methods:** A systematic review and meta-analysis was conducted on the association of endometriosis with anogenital distance. Three databases were queried in the identification phase, and a random-effects meta-analysis was applied to the data in studies that met the inclusion criteria.

**Results:** Shorter anogenital distance AF, measured from the anus to the posterior fourchette, was significantly associated with higher risk of endometriosis in the meta-analysis. By contrast, there was no such association for anogenital distance AC, measured from the anus to the clitoral surface. Both analyses demonstrated significant heterogeneity across studies. Too few studies were available for robust investigation of publication bias.

**Conclusions:** The association of short anogenital distance with endometriosis risk provides support for the hypothesis that endometriosis represents, in part, a disorder mediated by relatively low testosterone levels in early prenatal development. This conclusions has notable implications for understanding the causes and treatment of endometriosis.

## Background

Endometriosis, which has been reported in about 5-10% of reproductive-aged women, is characterized by endometrial tissue establishing outside of the uterus, most often in the ovaries, peritoneum, or rectovaginal region [1]. This ’ectopic’ endometrial tissue undergoes menstrual cycle changes similar to those of normally situated, ’eutopic’ endometrium, proliferating under the influence of estradiol. Endometriosis often causes severe pelvic pain, as well as reduced fertility in many patients. Management typically involves various means to reduce the effects of estrogenic stimuli that promote endometrial tissue proliferation, and surgery to remove endometriotic tissue [2,3].

Endometriosis risk is mediated by many genes each of small effect [4], exhibiting a heritability of approximately 0.50 [5,6]. GWA (genome wide association) studies have identified a set of single nucleotide polymorphisms (SNPs) that account for a small proportion of this heritability, and that includes SNPs in genes with demonstrated roles in early sexual development (WNT4, HOXC6), steroid hormone signalling (ESR1, GREB1, KDR), overall growth (IGF1), HPO axis function (FSHB), among other phenotypes [4]. Endometriosis risk is also mediated by environmental steroid effects, as exemplified by links of the disorder with the synthetic estrogen diethylstilbestrol, via influences during prenatal development [7,8].

A series of studies has tested for associations of endometriosis with anogenital distance (AGD), which represents an indicator of levels of prenatal testosterone in early fetal development (reviews in [9,10]). Anogenital distance, which is measured from the anus to landmarks on the genitalia, is substantially longer on average in males than in females among humans and other mammals, due to variation between the sexes in levels of prenatal testosterone that control growth of the perineal region. AGD also varies substantially within each sex, as indicated by observational studies and extensive experimentation with non-human animals, as well as studies of humans naturally subject to altered levels of testosterone [11–14]. Female mammals that develop under relatively low levels of prenatal testosterone thus tend to develop relatively short AGDs, and high levels result in longer AGDs [15].

Anogenital distance in women with endometriosis was initially studied in the context of developing new biomarkers for this disorder [16]. More recently, AGD has been analyzed in the context of the developmental basis of endometriosis, whereby relatively low prenatal testosterone differentially programs the developing hypothalmic-pituitary-ovarian (HPO) axis in such a way as to increase risk in later life [17,18]. This paradigm provides a novel conceptual and empirical framework for understanding and studying endometriosis, in a comparable way to analyses of the well-established role of relatively high prenatal testosterone in risk for development of polycystic ovary syndrome [17].

A suite of studies of anogenital distance in women with endometriosis, compared to controls, has been published to date. These studies all include the two main measures of AGD: AGD-AF (from the anus to the posterior fourchette), and AGD-AC (from the anus to the clitoral surface), and the results are heterogeneous both across studies and between these two measures of AGD. The purpose of this analysis is to conduct the first meta-analysis of AGD in relation to risk of endometriosis, to ascertain the strength of evidence overall, and to help guide future work.

## Methods

Three databases were searched systematically on 23 November 2023. First, Web of Science was searched using the search terms ’anogenital distance endometriosis’ in the ’All fields’ search box, which returned a total of 33 entries. Second, PubMed was searched using the search terms ’anogenital distance endometriosis’, which generated 21 returns. Third, Google Scholar, a more broadly inclusive search engine, was also searched using the terms ’anogenital distance endometriosis’, for any date, and with sorting by relevance. For this latter database, the first 100 citations were included in the initial screening. The returned articles were searched to determine if they met the search criteria (data on AGD from women with endometriosis, and from controls), and the other criteria for inclusion (unique data set not used in published work previously, and conduct of measurements for both AGD-AF and AGD-AC). A PRISMA diagram depicts the search and inclusion processes (Figure 1).

**Figure 1.**
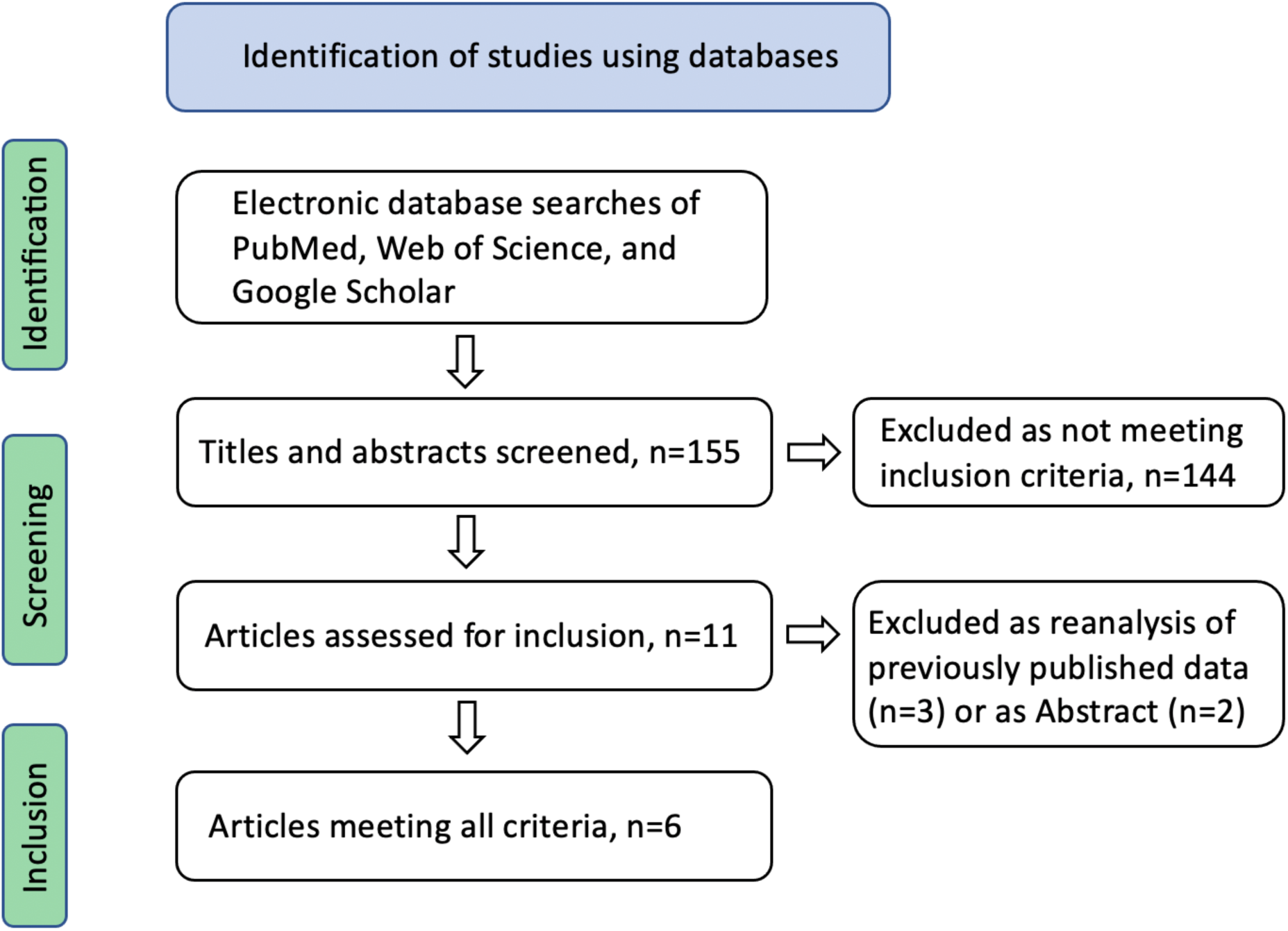
PRISMA diagram showing identification, screening, and inclusion process for the meta-analysis.

A random-effects model meta-analysis was conducted using the program Meta MAR [19]. The main goal of the meta-analysis was to increase statistical power to test for differences in AGD-AF, AGD-AC, or both, between women with endometriosis and controls. A random-effects model was used so that the results can be applied beyond the included studies, and given that women with different forms and severities of endometriosis may be subject to different magnitudes of effects on AGD (see e.g., [20]).

For the study [21] that included measurements of AGD-AF and AGD-AC from women with two different forms of endometriosis (ovarian and deep-infiltrating) separately but not together, the authors provided a copy of their data set, which allowed calculation and use of the data for all women with endometriosis combined.

## Results

A total of six studies met the inclusion criteria (Figure 1), comprising a total of 883 subjects overall, and were subject to meta-analyses for AGD-AF and AGD-AC. The random-effects meta-analysis demonstrated a significant difference between women with endometriosis and controls for AGD-AF (SMD=-0.77, p = 0.026, Figure 2). By contrast, the meta-analysis indicated no significant difference for AGD-AC (SMD=-0.29, p = 0.166, Figure 2). Both analyses demonstrated significant heterogeneity between studies (AGD-AF, Cochran’s Q = 47.4, P < 0.001; AGD-AC, Cochran’s Q = 30.8, p < 0.001). There was an insufficient number of studies available for robust interpretation of funnel plots in the context of testing for publication bias.

**Figure 2.**
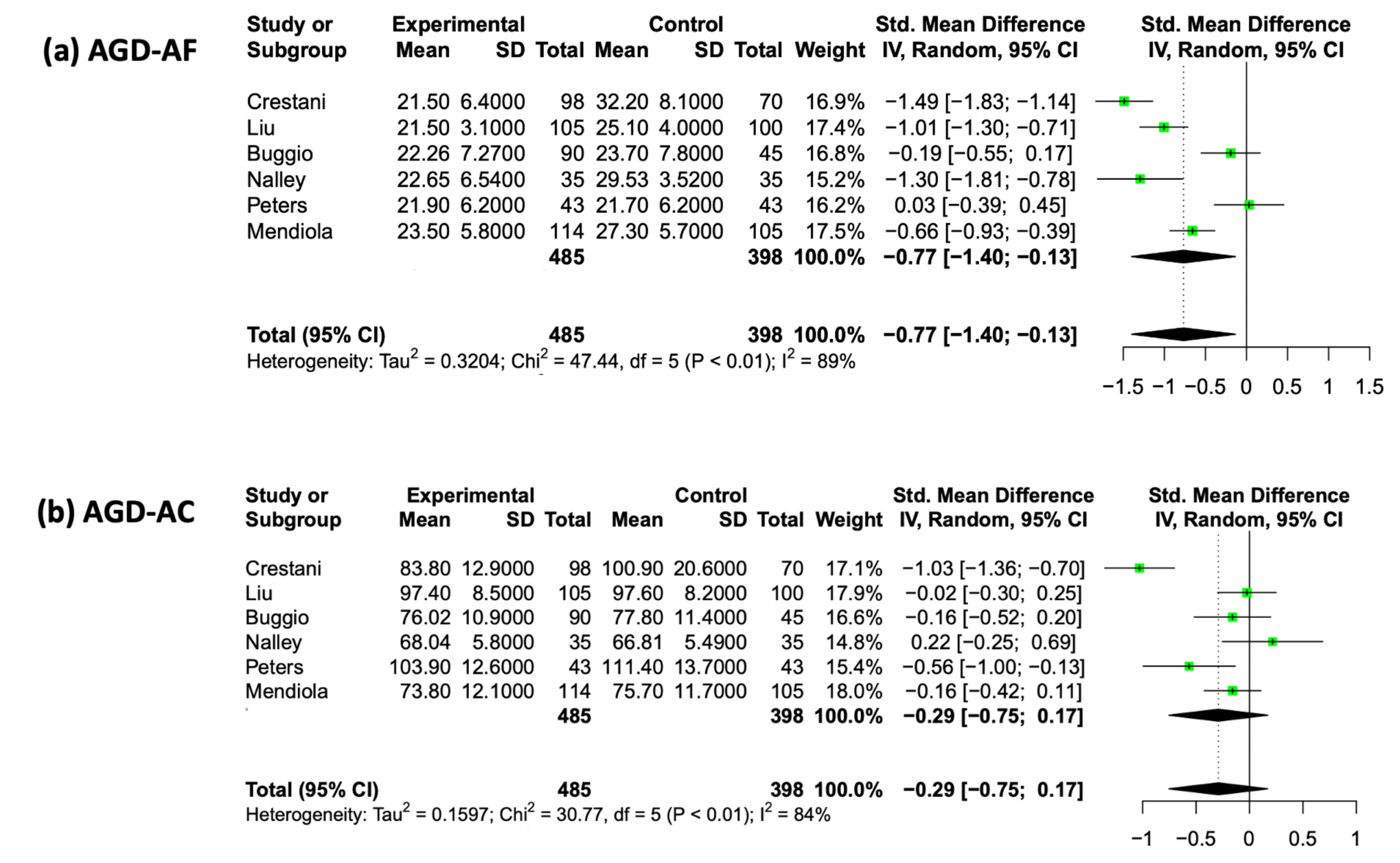
Main results of meta-analyses for (a) AGD-AF and (b) AGD-AF.

## Discussion

The primary finding of this study is that meta-analysis provides support for the hypothesis that AGD-AF is significantly shorter among women with endometriosis than among controls. By contrast, the comparison for AGD-AC was non-significant. The results for AGD-AF indicate that endometriosis shows a robustly replicated association with this measure of prenatal testosterone, such that low levels of this hormone in early prenatal development represent a risk factor, even if the predictive value of AGD-AF as a biomarker for diagnosis of endometriosis is limited [16,22]. These findings are of importance given that: (1) two studies have recently demonstrated that genetically predicted lower serum testosterone is significantly associated with higher risk of endometriosis [23,24], and (2) shorter anogenital distance (AGD-AF) has been linked with lower serum teststerone in healthy women of reproductive age [25].

The meta-analyses indicated significant heterogeneity among studies, for both AGD-AF and AGD-AC. Some of this variation is likely due to the populations analyzed; for example, the studies differed in the proportions of women with different forms of endometriosis (deep-infiltrating, ovarian, or peritoneal), in the body mass index (BMI) of participants, and in other variables. For example, nine of the 43 women with endometriosis in one sample [26] exhibited hyperandrogenism (and eight reported acne, and two showed hirsutism), which is characteristic of polycystic overy syndrome (PCOS), and may be related to the lack of difference in AGD-AF between women with endometriosis and controls in this study.

The differences in results reported here between AGD-AF and AGD-AC should be considered in the context of the AGD-AF distance being a subset of the AGD-AC distance, such that AGD-AC is equal to the sum of AGD-AF plus the distance between the posterior fourchette to the clitoral surface. As such, AGD-AF and AGD-AC are related as part-whole, and are positively correlated with one another, with correlation coefficients of between 0.33 and 0.60 (mean=0.48, N =7) [16,26–31]. Given that the overall average distance of AGD-AF is about one-third that of the overall average distance of AGD-AC, a substantial proportion of the mean difference between women with endometriosis and controls in AGD-AC can thus be attributed to the differences in AGD-AF. As such, for the results reported here, AGD-AF can be considered as the metric that differs significantly between women with endometriosis compared to controls, with any differences in AGD-AC (in some individual studies) being attributable at least in part to its variation. These inferences are demonstrated most clearly in the results from Mendiola et al. [16] and Buggio et al. [21], where the distance AGD-AF is 50–100% of that for AGD-AC, such that the former accounts for the bulk of the variation in the latter.

The biological significance of AGD-AF, compared to AGD-AC, is supported by the observations that: (1) the approximately two-fold difference between females and males for distance of the perineum (measured from the anus to the genitalia: posterior fourchette in females or base of scrotum in males), and corresponding to AGD-AF, is proportionally much larger than the sex difference in AGD-AC (measured in males from the anus to the anterior base of penis) (e.g., [32]); (2) among rodents, the longer AGD in males than females, which is known from experiments to be due to higher prenatal testosterone in males, corresponds to AGD-AF among humans; (3) AGD-AF, but not AGD-AC, is positively correlated with levels of serum testosterone in young women without endometriosis or PCOS [25,28]; and (4) AGD-AF is a better predictor of endometriosis than is AGD-AC in studies that use AUC (Area Under receiver operating Characteristic curve) analyses [27,33].

Significantly shorter AGDs (for AGD-AF, AGD-AC, or both) have also been associated with four correlates of endometriosis: premature ovarian insufficiency [34,35], dysmenorrhea [36,37], reduced responses to ovarian stimulation in IVF [29,38], and relatively regular menstrual cycles [39]. These findings support the hypothesis of causal links between lower prenatal testosterone and higher risk of endometriosis, as does evidence suggesting that women with endometriosis exhibit a higher level of sexual difference from males than do healthy women for a suite of sexually dimorphic traits [40]. Most broadly, these results provide evidence that endometriosis represents, in part, a reproductive disorder mediated by early development whereby relatively low testosterone affects the expression of many sexually dimorphic and sex limited traits [18]. This hypothesis is convergently supported by recent Mendelian Randomzation studies showing that lower genetically predicted testosterone is linked with higher risk of endometriosis, such that the relationship between the two shows evidence of causality [23,24]. Future studies that investigate effects of low prenatal testosterone in women, in the context of other correlates of endometriosis (such as early menarche and higher pain sensitivity), should provide further insights into the causes of this disorder. Studies testing for shorter AGDs in daughters of women with endometriosis, compared to controls, would also represent useful tests for effects of low prenatal testosterone in endometriosis risk.

The main limitations of this study include the relatively small number of studies conducted thus far on AGD in relation to endometriosis, the low sample sizes of some of the studies, their restriction thus far mainly to women of European descent, and the heterogeneity among studies in the clinical characteristics of the particpants. These limitations should be addressed with larger studies of more-diverse participants, and analyses of the genetic basis of AGD and its relationship to endometriosis risk factors.

In contrast to these studies of endometriosis and its correlates, analyses of PCOS provide evidence for longer AGDs in women with this disorder. Thus, in three data sets, women with PCOS showed significantly longer AGDs compared to controls [27,28] or to women with endometriosis [26], and in a fourth study, the difference between women with PCOS and controls was marginally non-significant (p=0.08 for AGD-AF, 0.17 for AGD-AC) [41]. Women with PCOS also show evidence of bearing daughters with longer AGDs, compared to controls, from three studies [42–44], whereas a fourth study showed no significant differences [45].

Longer AGDs in women with PCOS, and in their daughters, may be related in part to their higher body mass indices (BMIs) (women with PCOS had significantly higher BMIs than did controls in most studies of AGD [26,27,41,46]), given that: (1) BMI is higher among women with PCOS than in controls (e.g., [47]); (2) BMI is positively correlated with AGD among women [16,26,29,31, 33]; (3) maternal obesity is associated with longer AGD among daughters [48,49]; (4) AGD is positively correlated with serum testosterone [25,28]; and (5) BMI is positively associated with levels of testosterone, which represent a key correlate of PCOS [50]. Additional studies are needed of AGD in women with PCOS, and in daughters of women with PCOS, in the context of maternal BMI and testosterone, for sufficient data to be available for meta-analysis of AGD in this disorder.

## Conclusions

This meta-analysis provides evidence that risk of endometriosis is mediated by lower anogenital distance (specifically, AGD-AF), which suggests that lower testosterone in prenatal development increases risk of this disorder. Further studies are needed on AGD-AF in relation to the correlates and potential causes of endometriosis.

## Data Availability

Data used in the meta-analyses are available in Figure 2

## Abbreviations

AGD: anogenital distance
AGD-AC: anogenital distance, anus to clitoral surfac
AGD-AF: anogenital distance, anus to posterior fourchette
BMI: body mass index
GWA: genome-wide-association
PCOS: polycystic ovary syndrome
SNP: single nucleotide polymorphism

## Declarations

### Ethics approval and consent to participate

Not applicable

### Consent for publication

Not applicable

### Availability of data and materials

The data (means, standard deviations, and sample sizes) used in the meta-analysis were taken from the respective studies that were included in the systematic review, and are shown in Figure 1.

### Approvals

The author has seen and approved the manuscript.

### Authors’ contributions

BC conducted all of the research and manuscript preparation.

## Acknowledgements

I am grategul to L. Buggio for provision of information allowing calculation of AGD across all participants in their article [21].

